# The Effect of Repeated Mass Drug Administration on the Transmission of Yaws: A Genomic Epidemiology Study

**DOI:** 10.1101/2024.10.27.24316187

**Authors:** Amber Barton, Petra Pospíšilová, Camila G Beiras, Lucy N. John, Wendy Houinei, Lorenzo Giacani, David Šmajs, Michael Marks, Oriol Mitjà, Mathew A Beale, Nicholas R Thomson

**Affiliations:** Parasites and Microbes Programme, Wellcome Sanger Institute, Wellcome Genome Campus, Hinxton, Cambridgeshire, UK; The Department of Biology, Faculty of Medicine, Masaryk University, Brno, Czech Republic; Fight Infectious Diseases Foundation, Hospital Universitari Germans Trías i Pujol, Badalona, Spain; Universitat Autònoma de Barcelona, Bellaterra, Spain; National Department of Health, Port Moresby, Papua New Guinea; Department of Medicine, Division of Allergy and Infectious Diseases, University of Washington, Seattle, WA, USA; Department of Global Health, University of Washington, Seattle, WA, USA; Faculty of Infectious and Tropical Diseases, London School of Hygiene & Tropical Medicine, London, UK; Hospital for Tropical Diseases, University College London Hospital, London, UK; Division of Infection and Immunity, University College London, UK

**Author notes:** co-senior authors (Mathew A Beale, Nicholas R Thomson).

## Abstract

**Background:** Yaws, a neglected tropical disease caused by *Treponema pallidum* subspecies *pertenue*, has evaded eradication, in part due to a high proportion of asymptomatic cases. A cluster-randomised trial in a yaws endemic area compared repeated mass drug administration (MDA) with one round of MDA followed by targeted treatment. Repeated rounds of MDA reduced active and latent prevalence of yaws but led to emergence and spread of azithromycin resistance to three children. Here we aimed to finely delineate the dynamics of *T. pertenue* sub-lineages over the course of this trial.

**Methods:** We performed whole genome sequencing directly on DNA from 263 swabs collected during this trial, recovering 222 good-quality *T. pertenue* genomes. We examined the phylogenetic relationships between genomes linked to geospatial and patient metadata.

**Findings:** We identified 29 fine-scale sub-lineages of *T. pertenue*, of which ten were eliminated by MDA, whilst 13 persisted in the control arm, one in the experimental arm, two in both study arms, and three were first observed after commencing MDA. The two persistent sub-lineages had non-synonymous mutations in penicillin binding proteins. One of these sub-lineages evolved macrolide resistance (N=3), and was associated with lowered treponemal antibody levels (p = 0.004) and longer ulcer duration (p = 0.015). Despite the study taking place within a relatively small geographic area (Namatanai District, in the Island of New Ireland, Papua New Guinea) sub-lineages were geographically clustered, and older children were more likely to share sub-lineages (p = 6×10^−9^).

**Interpretation:** Our findings show that the re-emergence of yaws after MDA was attributed to multiple sub-lineages. The majority of these sub-lineages were detected in the population prior to MDA, and participants were more likely to share sub-lineages within the same ward, suggesting that re-emergence was mostly driven by local transmission. These findings could help inform future yaws elimination strategies.

**Funding:** European Research Council, European Union, Provincial Deputation of Barcelona, Barberà Solidaria Foundation and Wellcome.

**Research in context:** *Evidence before this study:* We searched PubMed on 24^th^ July 2024 using the terms treponema * pertenue OR yaws, genom* OR sequencing, and “mass drug administration” OR azithromycin OR “mass treatment”, without restrictions for language or date. Two studies were previously published on the current cohort from the Namatanai Province, Papua New Guinea, comparing three rounds of mass drug administration (MDA) with one round of MDA followed by targeted treatment. These studies used multi-locus sequence-typing and found that repeated MDA limited yaws to one sequence-type but resulted in three cases of macrolide resistance. A separate study used whole genome sequencing to find that after a single round of MDA on Lihir Island, Papua New Guinea, a rebound in yaws cases was caused by multiple sub-lineages of the same MLST, but evolution of macrolide resistance only occurred once. No studies have yet examined how repeated MDA affects the whole-genome diversity and evolution of yaws.

*Added value of this study:* Our findings show that re-emergence of yaws after MDA was caused by multiple sub-lineages, most of which were already present in the population before MDA. In the group undergoing three rounds of MDA there was still a small rebound in cases six months after the third round, caused by two “persistent” sub-lineages with mutations in penicillin-binding proteins. One of these sub-lineages was associated with lower treponemal antibody and developed macrolide resistance. Sub-lineages were more likely to be shared between older participants and those in close geographical proximity.

*Implications of all the available evidence:* These data suggest that re-emergence is predominantly driven by cases missed by the initial round of MDA rather than by importation of new cases. Much more efficient population suppression was achieved using three rounds of MDA, and this is likely due to more comprehensively treating the population and eliminating latent cases. Geographical clustering of sub-lineages suggests that elimination by maintaining cases at a low enough prevalence to result in stochastic “fade out” could be feasible and achieved by repeated mass drug administration. Transmission was found to be most common amongst older children, and targeted approaches focusing on these groups may be beneficial. However, ongoing surveillance for macrolide resistance will be needed to achieve eradication.

## Introduction

The burden of the neglected tropical disease yaws, caused by infection with *Treponema pallidum* subspecies *pertenue (T. pertenue)*^1^, is highest in Papua New Guinea and the Solomon Islands^2^. Transmission is thought to be driven by skin-to-skin contact, with 75% of new cases in those aged below 15 years^1^. While primary lesions heal in 3-6 months, disseminated secondary lesions affect skin and bones, and late-stage tertiary yaws can result in severe bone lesions^1^. Without an eradication campaign, yaws would cause an estimated 1·6 million disability-adjusted life years from 2015-2050^3^.

A worldwide elimination campaign led by WHO and UNICEF between 1952-1964^4^ successfully reduced yaws 95% by treating clinically apparent cases with benzathine benzylpenicillin (BPG) injections. However, due to logistical barriers in using BPG in remote locations, combined with a high prevalence of asymptomatic latent cases, yaws has evaded eradication. In 2012 a randomised controlled trial established that single-dose oral azithromycin had a 96% efficacy in treating yaws^5^, raising the possibility of mass drug administration (MDA). This prompted WHO to launch the Morges Strategy for yaws eradication, providing MDA in endemic communities followed by targeted treatment of clinical cases at six-month intervals^6^.

In 2013-2016, a longitudinal study on Lihir Island, Papua New Guinea, found that the Morges strategy only reduced yaws cases temporarily, before a recrudescence driven by three distinct sub-lineages of multi-locus sequence-type (MLST) JG8/J_E_11^7,8^. Furthermore, five epidemiologically-linked cases with the A2059G 23S rRNA macrolide resistance mutation were detected. 36-61% of cases identified during targeted treatment rounds were due to residents missing the initial MDA, suggesting that multiple rounds may be necessary to disrupt transmission. To evaluate this strategy, a 2018-2019 cluster-randomised trial comprising 56,676 individuals was carried out in the Namatanai District of the Island of New Ireland, Papua New Guinea, comparing a control group following the Morges strategy (one round of MDA followed by two rounds of active case treatment every six months) with an experimental group undergoing three rounds of MDA^9^. After 18 months, active cases were reduced by three-fold in the control group, and 11-fold in the experimental group. However, three epidemiologically linked cases harbouring the 23S rRNA A2058G macrolide resistance mutation were identified at the final timepoint in the experimental group^10^.

Whilst MLST indicated that molecular diversity fell after MDA, 93% of cases belonged to a single sequence type^9^. We hypothesised that genome-scale resolution would allow us to define sub-lineages within the same MLST, allowing us to trace their distribution over the study. We therefore performed direct whole genome sequencing (WGS) on samples from the Namatanai trial and confirmed that MDA had a dramatic impact on *T. pertenue* diversity, and that further rounds of MDA prevented the rebound in diversity seen in the control group. We further used these genomic data to investigate the potential fitness advantages of two sub-lineages which persisted in both arms throughout the study.

## Methods

### Study population

Study samples were collected during a cluster-randomised trial where 38 wards in three local-level governments areas (LLGs) were randomised to either a control group following the Morges strategy or an experimental group undergoing three rounds of MDA (appendix 1 p2)^9^. Estimated population coverage ranged from 65% in the second treatment round to 99% in the follow-up survey. The trial protocol was approved by the Medical Research Advisory Committee of the Papua New Guinea National Department of Health. Informed oral consent was obtained from all trial participants, or for children, the parents/guardians of trial participants.

### Whole genome sequencing and analysis

From 297 PCR-positive swabs, 263 unique samples underwent library preparation using the previously described pooled sequence-capture approach^8^ and whole genome sequencing (appendix 1 p2). Sequencing reads were prefiltered, trimmed and down-sampled then mapped to Samoa D *T. pertenue* reference genome (appendix 1 p2). We detected recombinogenic regions (appendix 1 p6), created a recombination-masked single nucleotide polymorphism (SNP) alignment including previously sequenced contextual samples, and inferred a maximum-likelihood phylogeny (appendix 1 pp2-3). Following ancestral reconstruction, samples were clustered into sub-lineages using rPinecone v0.1.0. with a SNP threshold of 1^11^. ARIBA version 2.14.6^12^ was used to infer MLST according to the scheme described by Medappa et. al^10^. Macrolide resistance alleles were inferred using the competitive mapping approach previously described^8^.

### Pair-wise comparisons

For every possible pair of samples where participant metadata was available, excluding comparisons between the same sample, we identified whether pairs fell into the same sub-lineage, whether they were from the same age group or sex, or whether they were from the same LLG, ward, or neighbouring wards (appendix 1 p3). We then assessed whether pairs sharing the same sub-lineage were over-represented in each of these categories using a chi-squared test. We calculated Bonferroni-corrected p-values to account for multiple comparisons.

## Results

### Namatanai *T. pertenue* genomes are genetically homogenous but can be subdivided into distinct sub-lineages

After sequencing and filtering the genomes for quality, our dataset comprised 222 novel *T. pertenue* genomes from Namatanai, and we contextualised these with 38 globally derived genomes, including 20 from nearby Lihir Island. Comparative analysis of genomes from Namatanai sequenced during this study indicated very close genomic relationships (Figure 1), with a median pair-wise SNP distance of 5 (range 0-139). Namatanai genomes fell within three phylogenetic clades corresponding to the MLST types J_E_11 (n = 204), S_E_22 (n = 11) and T_E_13 (n = 7) described elsewhere^9,10^ and corresponding to types JG8, SE7 and TD6 described previously^7,8^. Our use of whole genome sequencing enabled us to finely delineate these clades into 29 sub-lineages and 29 singletons with a distance of only 0-2 SNPs within each sub-lineage. For the purposes of this study, we adopted a sub-lineage naming system based on their properties within the trial (“PERS”-sub-lineage persisted in both arms throughout; “EXP”-sub-lineage found in the experimental group only; “ELIM”-sub-lineage eliminated from all trial groups; “ELIM_Exp”-sub-lineage eliminated from the experimental arm only; “EMERG”-sub-lineage first detected after Round 1 MDA).

**Figure 1:**
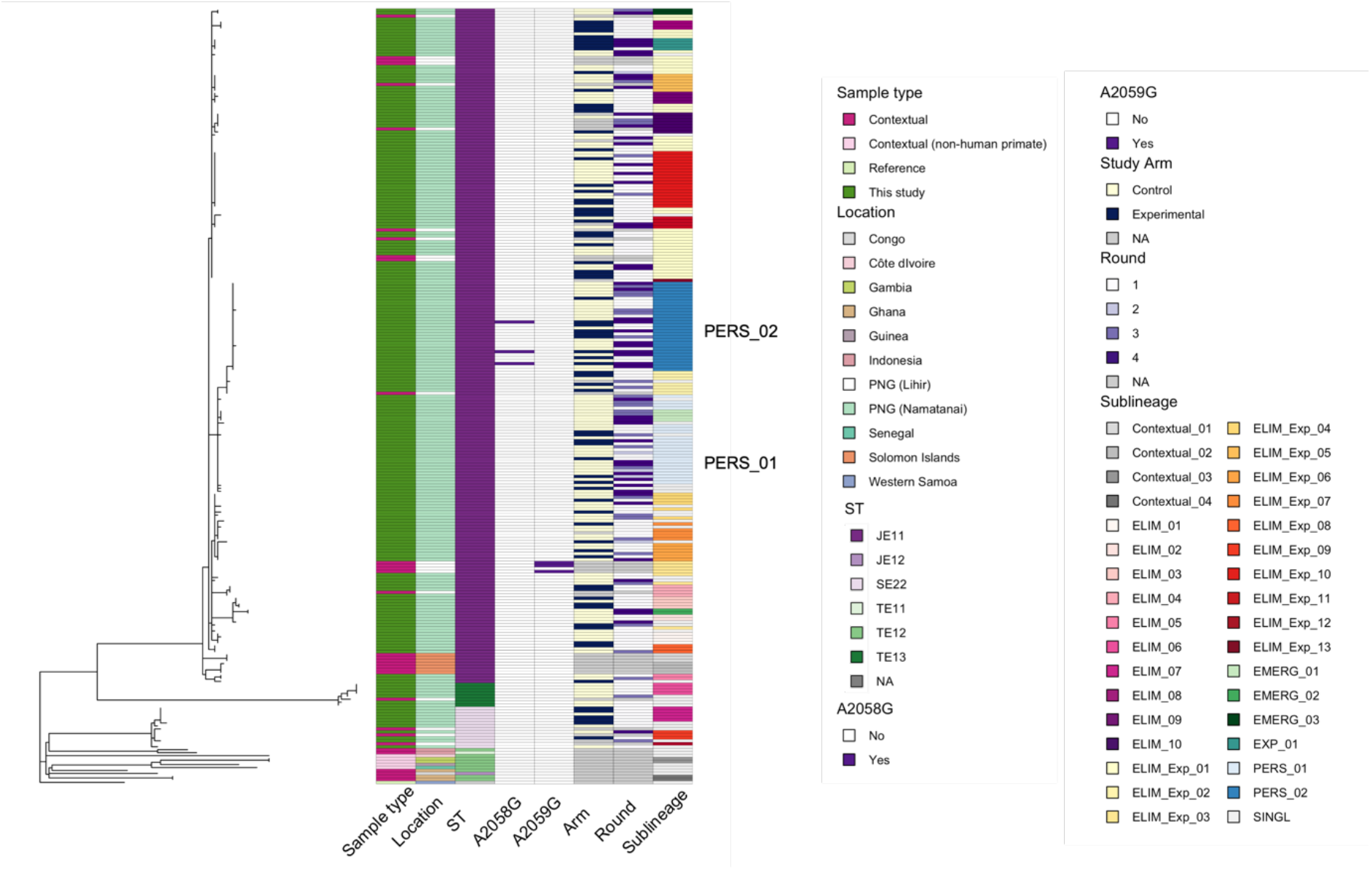
Maximum likelihood whole genome sequencing phylogeny for samples collected in Namatanai and contextual genomes. A heatmap indicates the sample type, location from which samples were collected, MLST type, presence of macrolide-resistance conferring mutations 23S rRNA A2058G and A2059G, study arm, study round, and sub-lineage as determined by rPinecone.

Our phylogenetic analysis using a globally derived genome collection indicated that genomes classified as ST J_E_11 from Papua New Guinea and the Solomon Islands were phylogenetically distinct from all other *T. pertenue* genomes (Figure 1 showing all samples analysed; appendix 1 p7 with the predominant Papua New Guinea J_E_11 clade collapsed). However, analysis within J_E_11 showed that although genomes from the Solomon Islands were phylogenetically distinct, those from Lihir Island, Papua New Guinea^8^ co-occurred in the same sub-lineages as those in this study, suggesting the two islands share a *T. pertenue* population. Moreover, although all *T. pertenue* strains harbouring macrolide resistance SNPs were classified as J_E_11 by MLST, the increased resolution afforded by WGS demonstrates that the sub-lineage that developed macrolide resistance through a 23S A2059G mutation in Lihir (sub-lineage ELIM_Exp_03 in Figure 1) was phylogenetically distinct from the sub-lineage which developed resistance through the A2058G mutation in Namatanai (PERS_02).

### *T. pertenue* genetic diversity was reduced after mass drug administration but rebounded within a year

We examined temporal patterns of *T. pertenue* genomic diversity, and observed a marked decline in Shannon’s diversity index in both study arms following the first round of MDA (from 2·9 to 0 in the control arm, and from 2·8 to 1·1 in the experimental arm; appendix 1 p8). Despite targeted treatment of clinical yaws cases, diversity in the control arm rebounded to near pre-MDA levels (2·7) within 12-18 months, in contrast to the experimental arm, where diversity remained low (1·0). To further delineate these patterns, we examined temporal changes in frequency for the 29 sub-lineages comprising two or more samples. Of these, ten sub-lineages observed at baseline were eliminated entirely during the study. A total of sixteen sub-lineages were detected at baseline and persisted after MDA, in the control arm (thirteen sub-lineages), the experimental arm (one sub-lineage) or both arms (two sub-lineages) (Figure 2a). Three sub-lineages were not detected at baseline but were observed after MDA in the control arm.

**Figure 2:**
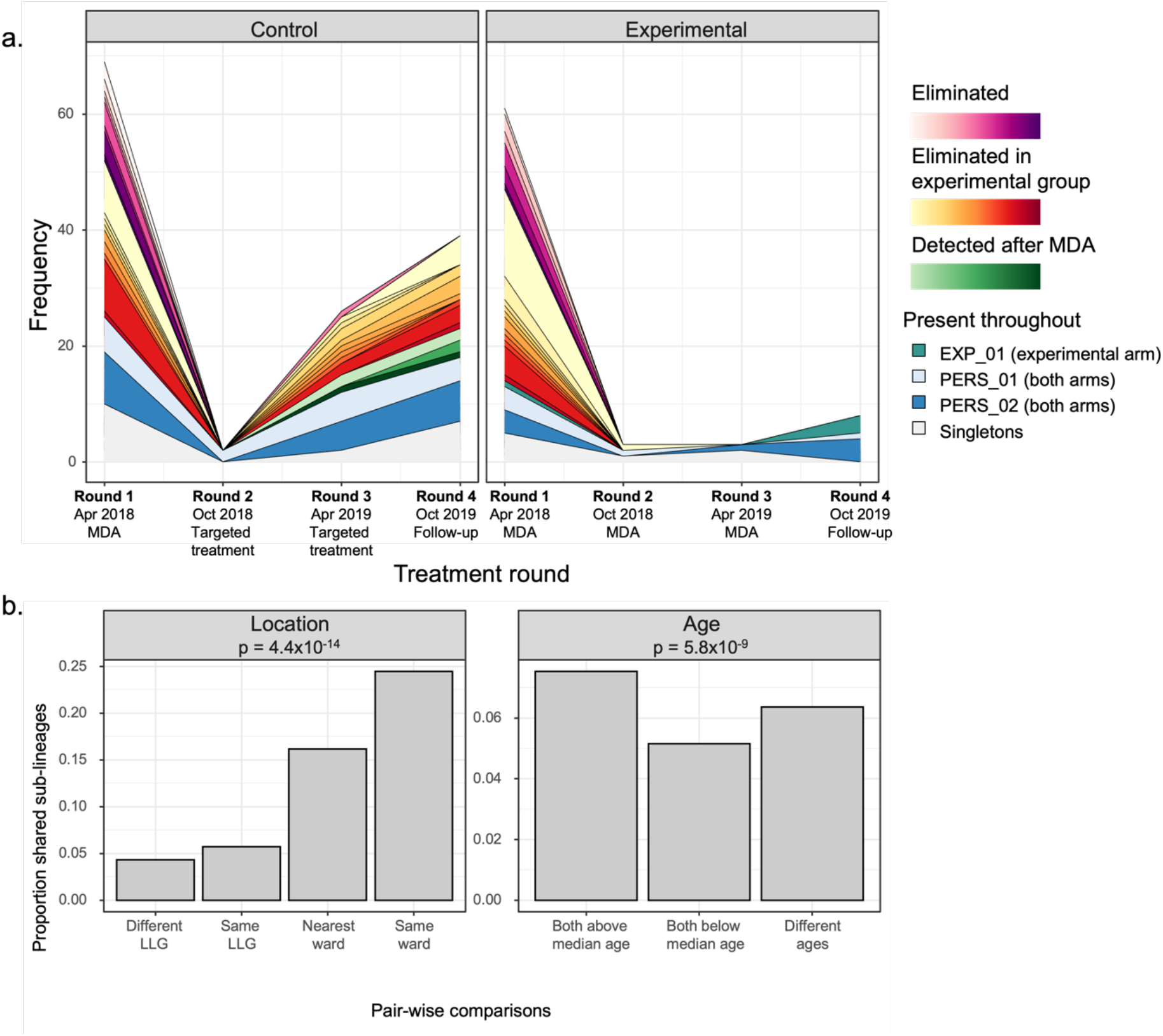
a. Frequency of different sub-lineages each treatment round, stratified by study arm. Sub-lineages that were eliminated entirely by MDA are shown in pink; those that were only eliminated in the experimental arm in orange; those that emerged in 2019 in green; those that were persistent throughout the study in blue, and singletons in grey. b. Proportion of pair-wise comparisons for which samples shared the same sub-lineage, stratified by sample location, study arm and age. Indicated p values are from a chi-squared test, Bonferroni-adjusted to account for each sample being compared to every other sample.

Next, we investigated whether recrudescence in the experimental arm was driven by individuals missed during MDA, transmission from the control arm, or importation. At the end of the study, eight samples representing three sub-lineages were present in the experimental group, of which two (PERS_01 and PERS_02) had been detected in both arms throughout. The single PERS_01 sample left in the experimental arm was both spatially and temporally closer to PERS_01 samples from the control arm compared with the experimental arm (appendix 1 p9). Three of the four PERS_02 samples had associated location metadata, and all three were in wards where PERS_02 had previously been detected (appendix 1 p10). However, they were also less than 10km away from control wards containing PERS_02 in the final round. Therefore, it was not possible to conclude whether PERS_02 persistence was driven by transmission from the control arm or within the experimental arm. EXP_01, which was only present in the experimental arm during the study, was detected in two wards in the final round, including the same ward it was originally detected in at baseline (appendix 1 p11). None of the sub-lineages present in the experimental arm in the final round had previously been found outside of the Namatanai District.

We next used pairwise comparisons to investigate geographical clustering of sub-lineages and found that pairs of samples from the same local-level government (LLG) were 1·3-fold more likely to share the same sub-lineage compared to pairs from different LLGs (Figure 2b). Likewise, pairs of samples from the same ward were 5.6-fold more likely to share the same sub-lineage compared to pairs from different LLGs, whilst pairs from neighbouring wards were 3.7-fold more likely. Notably in both the Namatanai and Sentral Niu Ailan LLGs, MDA had a dramatic effect on *T. pertenue* diversity, followed by a rebound in the control arm (appendix 1 p12). In contrast, *T. pertenue* diversity in the Matalai LLG was relatively low at the start of the study and remained relatively unaffected by MDA or targeted treatment, with the same two persistent sub-lineages dominating at the end of the study. This was still the case when the Namatanai and Sentral Niu Ailan LLG’s sample size was adjusted to match that of Matalai LLG, suggesting this pattern is not wholly attributed to the smaller population of the Matalai LLG (appendix 1 p13).

Finally, we examined whether demographic factors were risk factors for transmission. Sex was not significantly associated with sharing sub-lineages (appendix 1 p14; p = 1, chi-squared test). Since the age of participants was unevenly distributed, we split participants into two equally sized groups, and those above the median age for this study (aged 9-56 years) were more likely to share the same sub-lineage with one another (p = 6×10^−9^, chi-squared test) compared to samples from younger participants (aged 1-8 years).

### Two persistent sub-lineages are characterised by non-synonymous mutations in penicillin-binding proteins

As described earlier, two phylogenetically related sub-lineages (PERS_01 and PERS_02) persisted throughout interventions in both arms, suggesting they might possess a fitness advantage. We looked for phenotypic differences that may explain this and found that participants infected with PERS_02 reported a longer ulcer duration and had lower optical densities for a treponemal antibody assay compared with other sub-lineages (appendix 1 p19; Figure 3a). There was no significant difference in non-treponemal antibody (appendix 1 p15). We then tested whether any SNPs, including those highlighted as part of potential recombination blocks, were associated with these phenotypes. Across 222 samples from the Namatanai study, 88 SNPs were found in at least 5% of genomes (appendix 1 pp19-21). While none of these SNPs were associated with a longer ulcer duration, 55 SNPs in 44 genes were associated with decreased treponemal antibody (one-sided Mann-Whitney test, false discovery rate (fdr) after Benjamin-Hochberg multiple-testing correction < 0·05; Figure 3b). Amongst these 44 genes, proteins with signal peptides (15/44, 34%) were over-represented relative to the rest of the genome (142/1024, 14%) (p = 0·0008, Fisher’s test).

**Figure 3:**
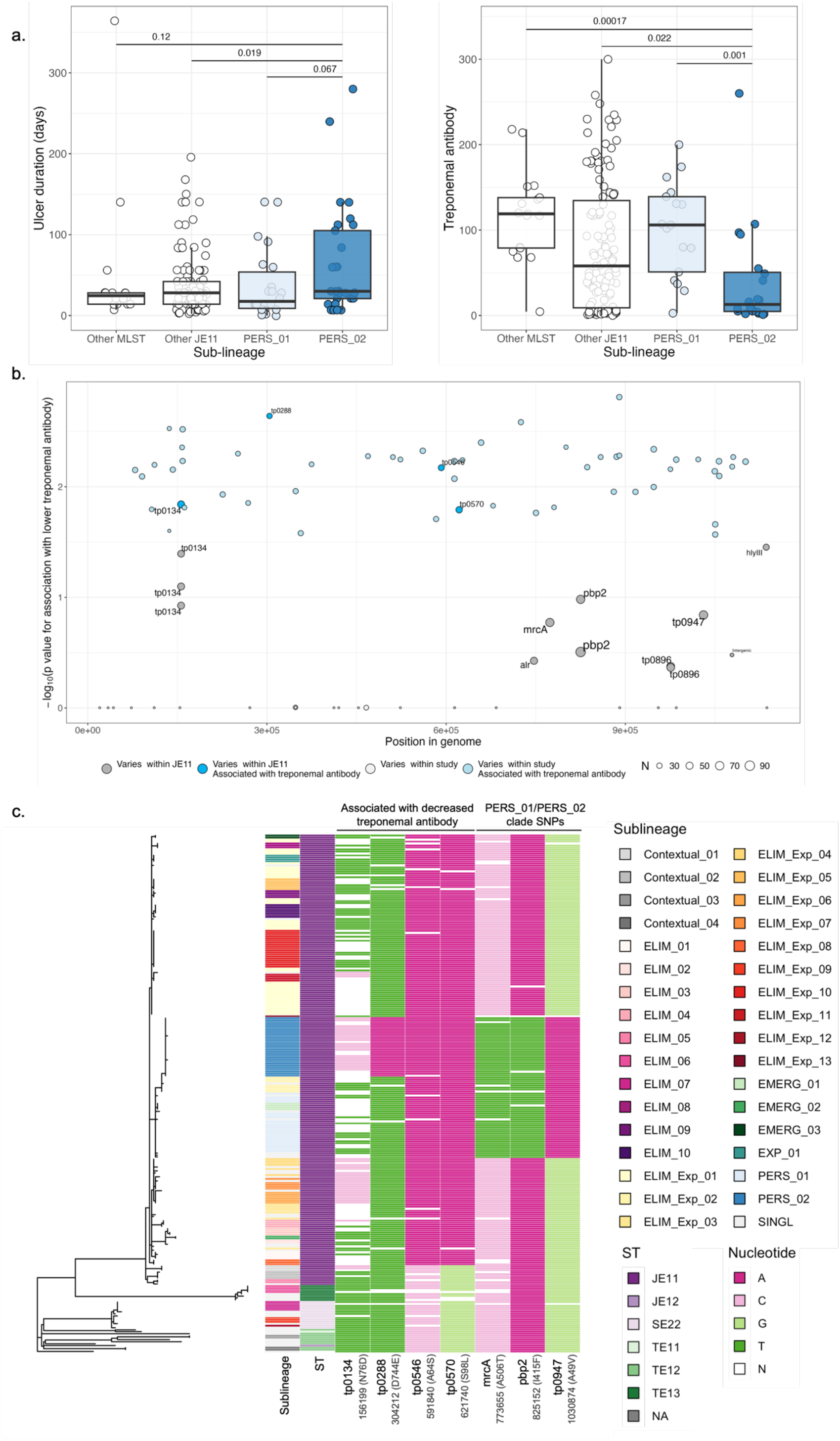
a. Optical densities for a treponemal antibody assay and reported ulcer duration in days in participants infected with PERS_01, PERS_02 and other sub-lineages. P values for a Mann-Whitney test comparing PERS_02 with each other group is indicated. b. Association of SNPs with lowered treponemal antibody compared with their position in the genome. SNPs with a minor allele frequency > 5% were included. SNPs are coloured by whether they were significantly associated with lowered treponemal antibody (one-tailed Mann-Whitney test, false discovery rate < 0·05) and whether they varied within the J_E_11 MLST. The size of each point reflects the frequency of the SNP. c. SNPs associated with lowered treponemal antibody that varied within J_E_11, and SNPs that were specific to the clade encompassing PERS_01 and PERS_02. For samples where no reads mapped to the SNP locus, the nucleotide is denoted as “N”.

Within the J_E_11 MLST, 16 SNPs were present in at least 5% of genomes and occurred along the ancestral phylogenetic branches differentiating J_E_11 sub-lineages from one another. Four of these SNPs were associated with decreased treponemal antibody (one-sided Mann-Whitney test, fdr < 0·05; Figures 3b and c), including a PERS_02-specific SNP causing a D744E mutation within the methyltransferase domain of cytoplasmic protein *tp0288*, which is involved with cell envelope biogenesis (appendix 1 p16). *Tp0134*, a paralogue of the adhesin *tp0136*^13^, varied at three sites within J_E_11 (G61S, D74E and N76D), of which N76D was associated with lower treponemal antibody, and ^13^differed between PERS_02 and PERS_01. Using the fine-scale whole-genome phylogeny previously generated in Beale et al. 2021^14^ (appendix 1 p17) we found that in *Treponema pallidum* subspecies *pallidum*, the causative agent of syphilis, *tp0134* varied at a site within the same loop (I78T) as *T. pertenue*.

Of the 16 SNPs which differentiated J_E_11 sub-lineages, three were specific to the clade encompassing PERS_01 and PERS_02 (Figure 3d). This included two non-synonymous mutations in penicillin binding proteins: I415F in the transpeptidase domain of *pbp2* (*tp0760*), and A506T in the transpeptidase domain of *mrcA* (*tp0705*) (appendix 1 p18). Both sub-lineages also possessed a A49V mutation in the cyclophilin-type peptidyl-prolyl cis-trans isomerase *tp0947*, an enzyme that catalyses an essential rate-limiting step in protein folding. We examined whether these SNPs were also present in *T. pallidum* (appendix 1 p17). I415F in *pbp2* was present in seven *T. pallidum* SS14-lineage samples in China, while A506T *mrcA* was globally distributed, having arisen in multiple clades within the *T. pallidum* Nichols (71/96 samples) and SS14 (30/407 samples) lineages. The two Nichols clades with A506T *mrcA* were the only clades within the Nichols lineage to develop macrolide resistance. Another variant at this site, A506V, was present in the globally distributed SS14 lineage (23/407). However, it is not known whether these SNPs confer an observable phenotype.

## Discussion

Understanding sources of yaws transmission will be key for informing intervention strategies if WHO is to achieve the target of yaws eradication by 2030^15^. In this study we used whole-genome sequencing to elucidate the dynamics and transmission of yaws in the Namatanai district following two possible MDA strategies. We observed that a single round of MDA only temporarily suppressed yaws diversity, followed by recrudescence mostly driven by sub-lineages that had been previously detected in the study population. Notably, two sub-lineages persisted in both arms of the study, of which one was associated with lower treponemal antibody and went on to develop three cases of macrolide resistance.

Inferring transmission of *T. pallidum* is challenging due to its high level of genome conservation, estimated to diverge by only one SNP every 6-8 years^14^. While MLST provides a useful tool for discriminating highly divergent genetic groups of *T. pallidum* at a global level^16^, it cannot distinguish sub-lineages at sufficient resolution to elucidate dynamics at a regional level. Previous research showed that *T. pertenue* in Namatanai was dominated by the J_E_11 ST, particularly following the first MDA round^9,10^. Here, the increased resolution afforded by whole-genome sequencing allowed us to subdivide samples considered identical by MLST into smaller sub-lineages with a distance of only 0-2 SNPs within each sub-lineage. Notably, we used the core genome here, and resolving and appropriately modelling incorporation of hypervariable regions using long-read or deep sequencing^17,18^ could enable further delineation of recent transmission. To maximise the value of our dataset we included genomes with up to 50% missing sites, reflecting the challenge of sequencing low-abundance pathogens. While it is possible that we missed SNPs at low quality sites that would further delineate sub-lineages, because our analysis accounts for missing sites along ancestral branches we do not expect this to substantially impact our conclusions.

We found that yaws diversity dropped after the first round of MDA, remaining low in the experimental arm following two further MDA rounds at six-month intervals, but rebounding over the same period in the control arm. Previously it was observed that after the first round of MDA, multiple *T. pertenue* MLST types were still present in the control arm, indicating that a single MDA does not represent a strong bottleneck^10^. Here we found that not only were multiple MLST groups present after MDA, but also multiple J_E_11 sub-lineages. 12/15 of these sub-lineages had been detected in the district before MDA, indicating that recrudescence was likely driven by a substantial number of cases missed during the initial round of MDA using the approach advocated by the Morges strategy. It is not yet clear whether diversity in the experimental group would return to similarly high levels 18 months after MDA cessation. However, repeated MDA successfully suppressed the *T. pertenue* population, which modelling suggests increases the likelihood of stochastic “fadeout”, where prevalence is low enough that stochastic effects can result in spontaneous eradication^19^.

Defining closely related but distinct sub-lineages also allowed us to quantify sub-lineage sharing between regions and sub-groups. Given the relatively small study region (covering a distance of approximately 300km) and slow genetic substitution rate of *T. pertenue*, we might have expected to find strains fully mixed throughout the region. However we observed substantial geographic clustering, which may be attributed to restricted travel options in these remote rural regions. This highly localised transmission suggests that yaws elimination could be more attainable than anticipated, as targeted treatment could be intensified in regions with clusters of cases, and clustering increases the likelihood of stochastic fadeout^19^. Spatial heterogeneity within implementation units will need to be considered when predicting prevalence. Furthermore, we observed sub-lineage sharing between neighbouring wards, indicating that implementation units should encompass at-least sub-districts in spatial scale. We also found that children aged nine or older, were more likely to share sub-lineages, which may reflect transmission chains within primary schools.

Finally, we investigated factors contributing to the success of the two sub-lineages that not only persisted but expanded in both study arms following MDA. These sub-lineages were both common at the beginning of the study, and harboured I415F and A506T mutations in penicillin-binding proteins *pbp2* and *mrcA* respectively. Analysing previous data, we found that the A506T *mrcA* variant also arose in both major syphilis lineages, while the I415F *pbp2* variant was present in syphilis samples from China^20^. To our knowledge this is the first time these variants have been reported in yaws. While it is possible these variants arose and expanded in both *T. pallidum* subspecies *pallidum* and *pertenue* due to random genetic drift, a more likely explanation is positive selection for these mutations. It is unclear whether they confer increased resistance to sub-therapeutic doses of penicillin, or some other fitness advantage, but if so, this may explain why these two sub-lineages were common at the beginning of the trial. A case-control study is currently underway to investigate whether these variants are associated with treatment failure in syphilis ^21^. Another possibility is that because these proteins are associated with cell envelope biogenesis, mutations could reduce growth rate *in vivo*. Not only could this extend the latency period, allowing *T. pertenue* to avoid targeted antibiotic treatment, but it could also enhance tolerance to antibiotics that interfere with growth^22^. Future confirmation of this association could inform decisions on whether to intensify treatment in regions where persistent sub-lineages are circulating.

Ultimately, only three cases in one of these two sub-lineages developed macrolide resistance over the course of the study (PERS_02). Those infected with this sub-lineage exhibited a longer ulcer duration and lower treponemal antibody levels. Proteins with signal peptides were over-represented amongst those with variants associated with reduced antibody, and PERS_02 possessed variants in membrane lipoprotein *tp0134*, a paralogue of the adhesin *tp0136*^13^. As periplasmic proteins are thought to be *T. pallidum’s* principal immunogens after killing by phagocytes^23^, variation in these proteins could prevent recognition. PERS_02 also harboured a mutation in a cytoplasmic protein involved in cell envelope biogenesis, which could slow bacterial growth and prevent *T. pallidum* reaching the critical capacity to activate immunity^23^. Previous studies have found that pathogens exhibit enhanced evolutionary rates within hosts compared to rates inferred at the population level^24–26^, suggesting that prolonged infections may increase the likelihood of *de novo* mutations such as those causing macrolide resistance. Prolonged infections also increase the likelihood of exposure to sub-therapeutic doses of azithromycin, creating a selection pressure for resistance.

Our findings here and elsewhere indicate that repeated MDA is more effective in reducing yaws cases than the single round MDA advocated in the Morges strategy^9^. However, this must be balanced against increased cost of conducting additional rounds of MDA. We observed marked geographic clustering even within a small area, suggesting that yaws elimination through targeting stochastic fade-out could be achievable. Furthermore, increased strain sharing between older children suggests that future campaigns targeting schools could offer a cost-effective solution for interrupting transmission. While three rounds of MDA did not achieve total elimination, repeated MDA did prevent resurgence in the diversity of yaws, and a strategy of long-term recurrent MDA on all neighbouring wards could be effective in maintaining prevalence at a sufficiently low level to achieve elimination. However, the potential selection pressure that repeated azithromycin MDA imposes towards macrolide resistance, both on *T. pertenue* and the microbiota, represents a reason for caution. Surveillance for macrolide resistance and treatment failure will be necessary for eradication. It is likely that a multipronged strategy involving MDA, targeted penicillin treatment and preventative interventions will be necessary to finally achieve eradication.

## Data Availability

All data produced are available online at under ENA project PRJEB40823.

https://www.ebi.ac.uk/ena/browser/view/PRJEB40823

## Contributors

AB and MAB carried out the data analysis. AB prepared the figures and wrote the original draft of the manuscript. LNJ and WH carried out the initial clinical study with support from CG-B and OM. PP and DS carried out laboratory work. AB, CG-B, MAB, NRT, OM, LG and MM interpreted the data. All authors reviewed and approved the final manuscript.

## Data sharing

Raw whole-genome sequencing data is deposited in the European Nucleotide Archive. Accession numbers and further data included in this analysis are provided in appendix 2.

## Acknowledgements

The cluster randomised trial was funded by Fundació “la Caixa”, Diputació de Barcelona, and Fundació Barberà Solidària. AB, MAB and NRT were supported by Wellcome funding to the Sanger Institute (220540/Z/20/A), and OM by the European Research Council under Grant Agreement number 850450. The work was partially financed by funds from the National Institute of Virology and Bacteriology project [Programme EXCELES, ID Project No. LX22NPO5103] funded by the European Union - Next Generation EU to DS.

The authors acknowledge support from the Sequencing teams, and Olivier Seret and the Parasites and Microbes Informatics team at the Wellcome Sanger Institute. This research was funded in whole, or in part, by the Wellcome Trust (220540/Z/20/A). For the purpose of open access, the authors have applied a CC-BY public copyright licence to any author-accepted manuscript version arising from this submission.

